# Determination of pathogenic nature of organisms isolated from aerobic respiratory culture/Multiplex PCR (Biofire® FilmArray) of lower respiratory tract samples in a tertiary care facility using a stepwise explorative model

**DOI:** 10.1101/2025.10.24.25338490

**Authors:** Shiv Narayan Sahu, Balram Ji Omar, Mukesh Bairwa, Prakhar Sharma, Partha Sahu, Prasan Kumar Panda

## Abstract

**Background:** Lower respiratory tract infections (LRTIs) remain a major cause of morbidity and mortality among hospitalized patients.^1^ However, isolating organisms from respiratory samples often leads to diagnostic uncertainty due to the coexistence of colonizers, commensals, and contaminants.^2^ To address this challenge, this study employed a structured, stepwise exploratory model to differentiate true pathogens from non-pathogens in aerobic respiratory cultures and Multiplex PCR (Biofire® FilmArray) results.

**Methods:** This prospective, longitudinal time-bound study was conducted over three months (August– October 2024) at a tertiary care center in northern India. Adult patients (≥ 18 years) with positive lower respiratory tract samples (aerobic culture or Multiplex PCR (Biofire® FilmArray) were enrolled. Each isolate was independently classified by the treating clinician, microbiologist, and study investigator using a six-step clinical-pathological algorithm that incorporated clinical signs, Sequential Organ Failure Assessment (SOFA) score trends, alternative infection sources, host factors, and outcome data. The final classification was determined by the investigator. Outcomes, including treatment response and mortality at 28 days, were compared across pathogen and non-pathogen groups.

**Findings:** Of the 145 included cases, 131 (90·3%) were classified as pathogens and 14 (9·7%) as non-pathogens. Cohen’s Kappa between investigator and microbiologist classifications was 0·28, indicating fair agreement. Among pathogen cases, 68 (51·9%) responded to treatment, while 63 (48·1%) did not respond to treatment in the pathogenic group. In contrast, 12 of 14 non-pathogen cases (85·7%) were not treated, with favourable outcomes in most, and only one unrelated death (7·1%).

**Interpretation:** The structured clinico -microbiological model strongly correlates with treatment outcomes, making it useful for differentiating infection from colonization. Crucially, microbiological detection alone doesn’t determine pathogenicity. Integrating clinical, laboratory, and outcome data is essential for rational antibiotic use and effective antimicrobial stewardship.

**Funding:** None

## Introduction

Lower respiratory tract infections (LRTIs) in admitted patients remain a leading cause of morbidity and mortality in hospitalized and critically ill patients worldwide.^1,3^ Clinical interpretation of positive respiratory cultures is often complicated by the coexistence of true pathogens, commensals, and colonizers.^2,4^ Misclassification can result in inappropriate antibiotic use, increased antimicrobial resistance (AMR), and unnecessary healthcare costs.^4^

Numerous studies have highlighted the pitfalls of equating microbial growth with infection, especially in ventilated or critically ill patients. Labelle et al. observed that 34% of patients with healthcare-associated pneumonia (HCAP) grew only colonizing flora, yet over 71% still received antibiotics, with no significant difference in mortality between culture-positive and culture-negative groups.^5^

The advent of rapid molecular diagnostics like multiplex PCR (e.g., Multiplex PCR (Biofire® FilmArray)® FilmArray) has improved organism detection and turnaround time, allowing clinicians to initiate early targeted therapy. However, these tests may also detect non-pathogenic flora or dead organisms, creating ambiguity in determining whether the identified microbe is responsible for active infection. Without clinical context, such high-sensitivity tools risk overdiagnosis and antibiotic overuse.^6,7^

While landmark studies support invasive sampling to guide antibiotic therapy, confirming the pathogenicity of isolated organisms remains challenging. In patients with prior antibiotic exposure, biofilm formation, or prolonged intubation, culture positivity may reflect colonization rather than true infection, raising concerns about over-reliance on microbiological results alone.^8,9^

This study aimed to differentiate true respiratory pathogens from non-pathogens (colonizers, commensals, and contaminants) in hospitalized adults using a stepwise explorative model that incorporates clinical features, SOFA score trends, risk factors for colonization, and outcome data. By correlating pathogen classification with treatment response and 28-day outcomes, this approach aims to improve diagnostic accuracy and antimicrobial stewardship in resource-limited, high-burden settings

## Methodology

This longitudinal explorative study was conducted to determine the proportion of pathogens and non-pathogens isolated from aerobic culture and multiplex PCR (Multiplex PCR (Biofire® FilmArray) of lower respiratory tract samples in hospitalized patients, using a stepwise exploratory model. The study was carried out at a 1000-bedded tertiary care teaching hospital in northern India (AIIMS Rishikesh) between August 1, 2024, and October 31, 2024. Data were collected from the Departments of General Medicine, Nephrology, and Pulmonary Medicine. The study received approval from the Institutional Ethics Committee. Adult patients aged 18 years or older admitted with a positive lower respiratory tract sample—either from conventional aerobic culture or Multiplex PCR (Biofire® FilmArray) —were eligible for inclusion. Patients were excluded if they declined consent, could not be followed up, or were excluded by the treating physician. Eligible patients were identified through daily visits to the Microbiology department. For each positive sample, the reporting microbiologist and the treating clinician were independently asked to classify the organism as either a pathogen or a non-pathogen, and their judgments were recorded. Baseline patient data, including vital signs, physical examination findings, laboratory parameters, and SOFA scores, were collected. Patients were followed until discharge or for a maximum of 28 days. Final classification of the organism was made by the study investigator, integrating microbiological data, clinical context, and patient outcomes.

### Structured stepwise model for organism classification (Fig. 1-2)

Whenever a culture or Biofire PCR tested positive and met inclusion criteria, the organism underwent a six-step clinico-microbiological algorithm to determine its pathogenicity by the investigator team as below. The patient is followed till discharge or 28 days, whichever is earlier, to determine the outcomes in the form of response to treatment, mortality and final characterization is done by the investigator.

### Flow chart (Figure 1)

#### Pathogen identification workflow (Supplementary)

- **Step 1:** Assess for any localized symptoms or signs of LRTI using **Checklist I** → If present → classify as pathogenic→ proceed to Step 4/5 → If absent → proceed to Step 2
- **Step 2:** Determine whether there is a rise in SOFA score by ≥ 2 points within 48 hours (**Checklist II**) → If yes → proceed to Step 3 → If no → classify as **Non-P athogenic** → proceed to Step 4/5/6
- **Step 3:** Is there any alternative source of infection other than LRTI (**Checklist III**)? → If no → classify as **Pathogenic** → If yes → classify as **Non-P athogenic** → proceed to Step 4/5/6
- **Steps 4, 5 & 6:** Characterization of Pathogenic/Non-Pathogenic organisms
- **Step 4:** Evaluate if the organism is part of normal flora (**Checklist IV**) → If yes → classify as Pathogenic Commensal/Non-pathogenic commensal → If no → classify as **Pathogenic-Direct** (directly pathogenic)
- **Step 5:** Are there host-related factors that support colonization (endotracheal tube/tracheostomy) (**Checklist V**) → If yes → classify as **Pathogenic Colonizer/Non-P athogenic Colonizer** → If no → classify as **D irect pathogen**
- **Step 6:** For non-pathogenic cases → If likely due to contamination (**Checklist VI**) → classify as **Non-pathogenic Contaminant**

### ALT Text

Flowchart illustrating the six -step clinical-pathological algorithm used for classifying organisms as pathogenic or non-pathogenic in lower respiratory tract samples.

*Note:* The Sequential Organ Failure Assessment (SOFA) score was calculated using standard criteria based on six organ systems: Respiratory, cardiovascular, hepatic, coagulation, renal, and neurological parameters, as per established guidelines.

### Outcome measures

The primary outcome measure was to determine the proportion of pathogens and non-pathogens in aerobic respiratory culture/Multiplex PCR (Biofire® FilmArray) in a stepwise model.

The secondary outcome measure was to determine the identified organisms as pathogenic colonizers, non-pathogenic contaminants, or non-pathogenic colonizers.

Once the pathogenicity of the isolated organism was determined, patients were followed prospectively until discharge or up to 28 days—whichever occurred earlier—to evaluate clinical outcomes. Based on the clinical course and treatment decisions, outcomes were classified into four categories: (1) Improved with treatment, applicable to patients with pathogenic organisms who showed clinical recovery following targeted antimicrobial therapy; (2) No response to treatment, representing pathogenic cases where appropriate therapy failed to produce clinical improvement; (3) Not treated, defined for non-pathogenic isolates where no specific antimicrobial therapy was initiated; and (4) Treatment initiated for non-pathogenic organisms. This categorization allowed objective assessment of the appropriateness of pathogen classification and its correlation with patient outcomes.

### Statistical analysis

Data entry was performed using Microsoft Excel, and statistical analysis was conducted using IBM SPSS Statistics for Windows, Version 25.0 (IBM Corp., Armonk, NY, USA). The collected data were analysed using a combination of descriptive and inferential statistical techniques. Continuous variables were expressed as mean ± standard deviation (SD) or median with interquartile range (IQR), depending on the distribution assessed. The normality of continuous variables was verified using visual and statistical methods, following which either *independent samples t-tests* (for normally distributed variables) or *Wilcoxon-Mann-Whitney U tests* (for non-parametric comparisons) were employed to assess group differences. Categorical variables were summarized as frequencies and percentages, and comparisons between groups were performed using the *Chi-squared test* or *Fisher’s exact test*, as appropriate. To assess agreement between categorical variables, particularly in organism classification by microbiologists versus investigators, *Cohen’s Kappa statistic* was calculated. The strength of associations was further evaluated using effect size metrics such as *Cramer’s V, Bias-corrected Cramer’s V*, and *point-biserial correlation* for appropriate pairings. A p-value of <0.05 was considered statistically significant for all tests.

## Results

A total of 145 culture-positive or positive Multiplex PCR (Biofire® FilmArray) PCR events were included in the study and evaluated using the structured six -step clinico-microbiological algorithm (Fig. 1). Each isolate was independently assessed by the microbiologist, treating physician, and the investigator for its pathogenic potential, incorporating clinical indicators, SOFA score dynamics, treatment response, and presence of foreign devices.

### Baseline characteristics

The mean age of participants was 50.4 ± 16.3 years, with 55.9% males. Most were from Uttar Pradesh (47·6%) or Uttarakhand (46·9%), and nearly half were admitted to the Medicine ICU. Aerobic culture was the predominant identification method (90·3%), with ET/TT as the most common sample type (66·2%). *Acinetobacter baumannii* (46·9%) and *Klebsiella pneumoniae* (42·1%) were frequently isolated. A majority had monomicrobial infections (75·9%), localized signs (95·2%), and elevated SOFA scores (76·6%) (Table 1). Common comorbidities included hypertension (45·1%), and diabetes (41·8%).

**Table 1.** Association of demographic, microbiologic, and clinical variables associated with pathogen vs non-pathogen.

|  |  |  |  |  |  |
| --- | --- | --- | --- | --- | --- |
| Category | Parameter | Total (N = 205) | Non-Pathogen (n=14 ) | Pathogen (n=131) | p-value |
| Demographic Variables | Age (Years) | 50.42 ± 16.35 | 51.21 ± 16.43 | 50.34 ± 16.40 | 0.923<sup>1</sup> |
| Gender | Male | 81 (55.9%) | 10 (71.4%) | 71 (54.2%) | 0.217<sup>2</sup> |
|  | Female | 64 (44.1%) | 4 (28.6%) | 60 (45.8%) |  |
| Site of clinical care | General Medicine Ward | 20 (13.8%) | 5 (35.7%) | 15 (11.5%) | 0.094<sup>3</sup> |
|  | Medicine ICU | 71 (49.0%) | 4 (28.6%) | 67 (51.1%) |  |
|  | Nephrology Ward | 5 (3.4%) | 0 (0.0%) | 5 (3.8%) |  |
|  | Pulmonary Medicine | 49 (33.8%) | 5 (35.7%) | 44 (33.6%) |  |
| Microbiological variables |  |  |  |  |  |
| Sample | Aerobic culture | 131 (90.3%) | 14 (100.0%) | 117 (89.3%) | 0.362<sup>3</sup> |
|  | Multiplex PCR (Biofire® FilmArray) | 14 (9.7%) | 0 (0.0%) | 14 (10.7%) |  |
|  | Sputum | 24 (16.6%) | 4 (28.6%) | 20 (15.3%) |  |
|  | ET/TT | 96 (66.2%) | 8 (57.1%) | 88 (67.2%) |  |
|  | BAL | 25 (17.2%) | 2 (14.3%) | 23 (17.6%) |  |
| Organism | *Klebsiella Pneumoniae* | 61 (42.1%) | 5 (35.7%) | 56 (42.7%) | 0.612<sup>2</sup> |
|  | *Acinetobacter Baumannii* | 68 (46.9%) | 4 (28.6%) | 64 (48.9%) | 0.148<sup>2</sup> |
|  | *Pseudomonas Aeruginosa* | 32 (22.1%) | 2 (14.3%) | 30 (22.9%) | 0.735<sup>3</sup> |
|  | Influenza Virus | 4 (2.8%) | 0 (0.0%) | 4 (3.1%) | 1.000<sup>3</sup> |
| Number of Organisms Positive/sample | 1 | 110 (75.9%) | 12 (85.7%) | 98 (74.8%) |  |
|  | 2 | 27 (18.6%) | 2 (14.3%) | 25 (19.1%) |  |
|  | 3 | 3 (2.1%) | 0 (0.0%) | 3 (2.3%) |  |
|  | 4 | 4 (2.8%) | 0 (0.0%) | 4 (3.1%) |  |
|  | 6 | 1 (0.7%) | 0 (0.0%) | 1 (0.8%) |  |
|  | Same Organism Isolated from the Local Site and Blood | 28 (19.3%) | 0 (0.0%) | 28 (21.4%) |  |
|  | Pathogen |  | 1 (7.1%) | 131 (100.0%) |  |
| Clinical Variables | Signs and Symptoms of Any Localized Site of Infection *** | 138 (95.2%) | 9 (64.3%) | 129 (98.5%) | <0.001<sup>3</sup> |
|  | Fever | 71 (51.4%) | 4 (44.4%) | 67 (51.9%) | 0.740<sup>3</sup> |
|  | Cough | 88 (63.8%) | 7 (77.8%) | 81 (62.8%) | 0.487<sup>3</sup> |
|  | Expectoration *** | 105 (76.1%) | 4 (44.4%) | 101 (78.3%) | 0.036<sup>3</sup> |
|  | Increasing Oxygen Requirement *** | 107 (77.5%) | 3 (33.3%) | 104 (80.6%) | 0.004<sup>3</sup> |
|  | Tachypnoea *** | 97 (70.3%) | 3 (33.3%) | 94 (72.9%) | 0.020<sup>3</sup> |
|  | Tachycardia | 68 (49.3%) | 2 (22.2%) | 66 (51.2%) | 0.166<sup>3</sup> |
|  | Chest Pain | 4 (2.9%) | 1 (11.1%) | 3 (2.3%) | 0.239<sup>3</sup> |
| | Increase in SOFA Score $\geq 2$ (48 Hours) *** | 111 (76.6%) | 3 (21.4%) | 108 (82.4%) | <0.001<sup>3</sup> |
| **Risk Factors for Colonization** | Invasive Mechanical Ventilation | 103 (71.0%) | 7 (50.0%) | 96 (73.3%) | 0.116<sup>3</sup> |
|  | Recent Hospitalization or Antibiotic Use Within the Past 90 Days | 86 (59.3%) | 8 (57.1%) | 78 (59.5%) | 0.862<sup>2</sup> |
|  | Poor Oral Hygiene Due to Tobacco Chewing, Smoking, Alcoholism, Dry Mouth, or Chronic Bedridden Status | 56 (38.6%) | 5 (35.7%) | 51 (38.9%) | 0.814<sup>2</sup> |
|  | Chronic Obstructive Pulmonary Disease (COPD) | 31 (21.4%) | 4 (28.6%) | 27 (20.6%) | 0.499<sup>3</sup> |
|  | Tracheostomy | 25 (17.2%) | 1 (7.1%) | 24 (18.3%) | 0.465<sup>3</sup> |
|  | Prior Isolation of the Organism Within One Year of the Incident | 22 (15.2%) | 0 (0.0%) | 22 (16.8%) | 0.129<sup>3</sup> |
|  | History of Lower Respiratory Tract Infection | 10 (6.9%) | 2 (14.3%) | 8 (6.1%) | 0.249<sup>3</sup> |
|  | Neuromuscular Disorders | 7 (4.8%) | 0 (0.0%) | 7 (5.3%) | 1.000<sup>3</sup> |
|  | Bronchiectasis | 3 (2.1%) | 1 (7.1%) | 2 (1.5%) | 0.264<sup>3</sup> |
| **Complications** | MODS (Multiple Organ Dysfunction Syndrome) | 71 (49.0%) | 4 (28.6%) | 67 (51.1%) | 0.108<sup>2</sup> |
|  | Ventilator-associated Pneumonia *** | 43 (29.7%) | 2 (14.3%) | 55 (42.0%) | 0.044<sup>2</sup> |
|  | Shock *** | 51 (35.2%) | 0 (0.0%) | 51 (38.9%) | 0.002<sup>3</sup> |
|  | Bloodstream Infections | 48 (33.1%) | 2 (14.3%) | 46 (35.1%) | 0.143<sup>3</sup> |
|  | Acute Kidney Injury | 16 (11.0%) | 1 (7.1%) | 15 (11.5%) | 1.000<sup>3</sup> |
|  | Patients on Hemodialysis | 21 (14.5%) | 0 (0.0%) | 21 (16.0%) | 0.222<sup>3</sup> |
|  | Arrhythmias, Including Ventricular Tachycardia and Fibrillation | 5 (3.4%) | 0 (0.0%) | 5 (3.8%) | 1.000<sup>3</sup> |
|  | Bedsore | 7 (4.8%) | 0 (0.0%) | 7 (5.3%) | 1.000<sup>3</sup> |
| **Comorbidities** | Diabetes | 38 (41.8%) | 3 (42.9%) | 35 (41.7%) | 1.000<sup>3</sup> |
|  | Hypertension | 41 (45.1%) | 2 (28.6%) | 39 (46.4%) | 0.451<sup>3</sup> |
|  | COPD (Chronic Obstructive Pulmonary Disease) | 26 (28.6%) | 3 (42.9%) | 23 (27.4%) | 0.403<sup>3</sup> |
|  | CAD (coronary artery disease) | 6 (6.6%) | 0 (0.0%) | 6 (7.1%) | 1.000<sup>3</sup> |
|  | CLD (Chronic Liver Disease) | 3 (3.3%) | 1 (14.3%) | 2 (2.4%) | 0.216<sup>3</sup> |
|  | CKD (chronic kidney disease) | 20 (22.0%) | 0 (0.0%) | 20 (23.8%) | 0.340<sup>3</sup> |
|  | Malignancy | 5 (5.5%) | 0 (0.0%) | 5 (6.0%) | 1.000<sup>3</sup> |

\*\*\*Significant at $p < 0.05$ , 1: Wilcoxon-Mann-Whitney U Test, 2: Chi-Squared Test, 3: Fisher's Exact Test
ALT Text: Statistical comparison of patient characteristics, risk factors, complications, and comorbidities between pathogen and non-pathogen groups.

### Outcomes

Among 145 respiratory samples, characterization by the investigator classified 131 (90·3%) isolates as *pathogens* and 14 (9·7%) as *non-pathogens*. A comparison between the investigators and the microbiologist’s classification of organisms as *pathogen* or *non-pathogen* revealed a Cohen’s Kappa coefficient of 0·28, indicating fair agreement between the two raters. The cross-tabulation shows that both parties agreed on 129 samples being pathogens, while agreement on non-pathogens occurred in only 3 cases (Table 2). The discordant was noted in 13 cases. The majority of disagreements occurred when the microbiologist labelled the isolate as pathogenic (based on culture identification), while the investigator labelled it as non-pathogenic. The investigator’s decision was based on a lack of overt clinical features of infection, which outweighed the observation of a rise in SOFA score that otherwise suggested organ dysfunction. (Table 3)

**Table 2.**
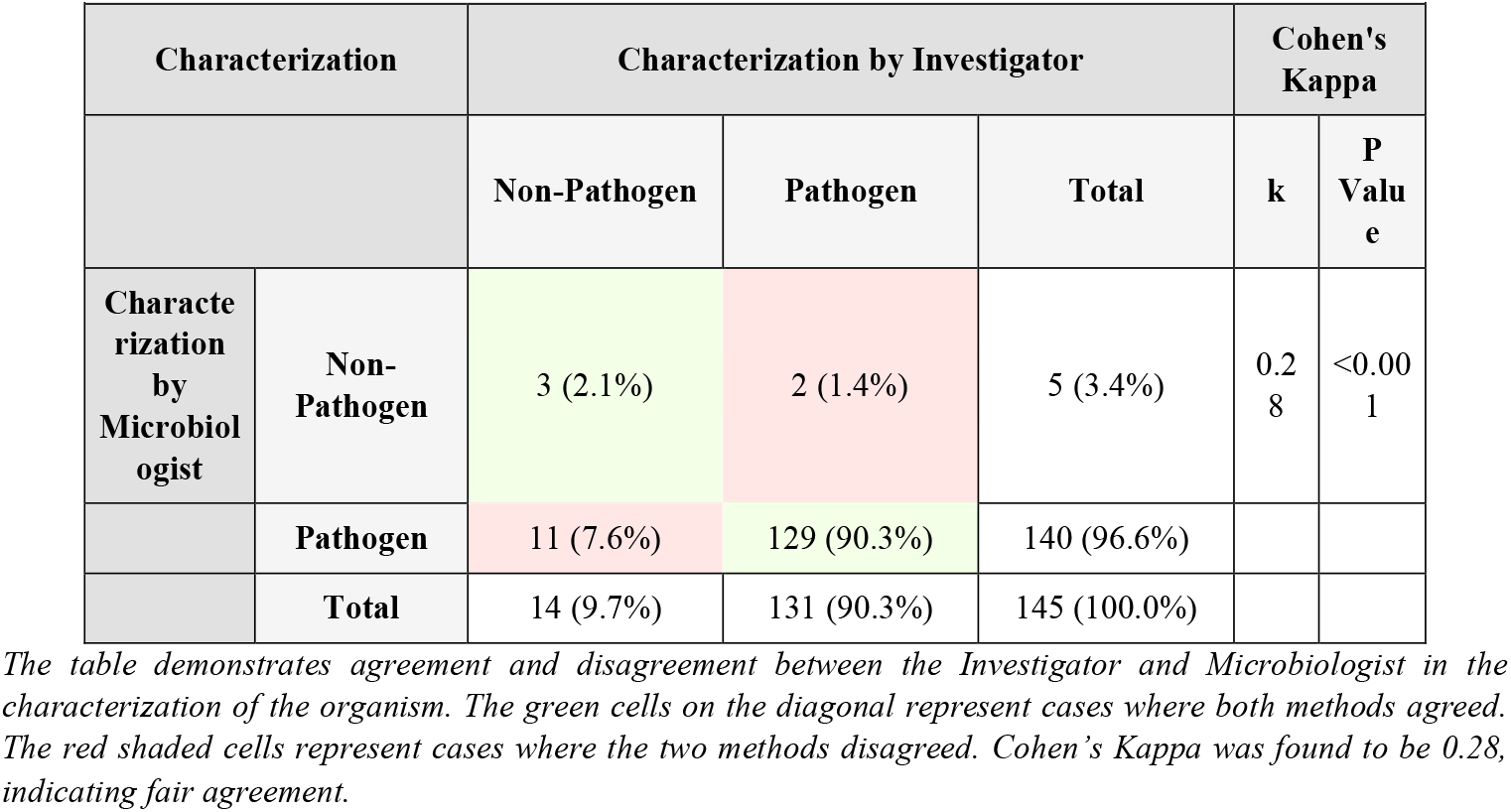
Agreement for Pathogen Characterization between Investigator and Microbiologist.

|  |  |  |  |  |  |  |
| --- | --- | --- | --- | --- | --- | --- |
| Characterization |  | Characterization by Investigator |  |  | Cohen's Kappa |  |
|  |  | Non-Pathogen | Pathogen | Total | k | P Value |
| Characterization by Microbiologist | Non-Pathogen | 3 (2.1%) | 2 (1.4%) | 5 (3.4%) | 0.28 | <0.001 |
|  | Pathogen | 11 (7.6%) | 129 (90.3%) | 140 (96.6%) |  |  |
|  | Total | 14 (9.7%) | 131 (90.3%) | 145 (100.0%) |  |  |

The table demonstrates agreement and disagreement between the Investigator and Microbiologist in the characterization of the organism. The green cells on the diagonal represent cases where both methods agreed. The red shaded cells represent cases where the two methods disagreed. Cohen's Kappa was found to be 0.28, indicating fair agreement.

The mean age of patients was comparable between groups (p = 0·923). While gender and state distribution did not significantly differ. Significantly fewer non-pathogen cases had signs of localized infection (64·3% vs 98·5%, *p* < 0 001), lower respiratory tract localization (57·1% vs 96·9%, *p* < 0.001), and SOFA ≥2 (21·4% vs 82·4%, *p* < 0 001). Organism distribution (e.g., *Klebsiella pneumoniae, Acinetobacter baumannii*) showed no significant variation between groups.

A significantly greater number of pathogen cases had respiratory symptoms such as expectoration (*p* = 0.036), increasing oxygen requirements (*p* = 0.004), tachypnoea (*p* = 0.020), and altered mental status (*p* = 0.011). Ventilator-associated pneumonia (*p* = 0.044), and septic shock (*p* = 0.011) were significantly more common in pathogen cases. (Table 1)

Out of 14 cases classified as non-pathogenic, 12 patients (85·7%) were not treated, and 2 patients (14 3%) were treated but remained clinically stable. None showed improvement attributable to antimicrobial therapy. One patient (7·1%) in this group died without receiving treatment; however, the mortality was attributed to ventricular arrhythmia and was unrelated to sepsis. Among 131 cases classified as pathogenic, 68 patients (51·9%) showed clinical improvement with treatment, while 63 patients (48·1%) showed no response despite therapy. A total of 48 patients (36·6%) in the pathogen group died, all of whom had failed to respond to treatment. Only 1 death (0·8%) occurred among those who improved with therapy secondary to another infection (ventilator-associated pneumonia). (Table 4)

Among 145 respiratory isolates, pathogenic colonizers were most frequent (n=103, 71.0%), followed by pathogenic direct pathogens (n=25, 17.2%) and pathogenic commensals (n=3, 2.1%), while non-pathogenic isolates comprised commensals (n=1, 0.7%), colonizers (n=5, 3.4%), and contaminants (n=8, 5.5%). The mean age and gender distribution were comparable across groups. However, sample type distribution differed significantly (p<0.001), with sputum predominant among direct pathogens and endotracheal aspirates among colonizers. Fever, cough, and increasing oxygen requirement were significantly more common in pathogenic isolates (p<0.001). Pathogenic colonizers exhibited the highest proportion of patients with SOFA score ≥2 (85.4%), shock (43.7%), and VAP (48.5%). Risk factors such as mechanical ventilation (93.2%) and recent antibiotic exposure (69.9%) were strongly associated with colonizers (p<0.001). Treatment response was best among pathogenic commensals (100%) and direct pathogens (80%), while most non-pathogenic isolates were either untreated or clinically stable. Mortality was highest among pathogenic colonizers (50.6%) compared with direct pathogens (16%) and non-pathogenic groups (0%) (p=0.001) (Table. 4).

## Discussion

This study addresses a critical diagnostic challenge in respiratory medicine: differentiating true infection from colonization or contamination in lower respiratory tract samples. By utilizing a structured, stepwise clinico-microbiological model, our research bridges the gap between microbiological identification and clinical relevance, particularly in high-burden, resource-constrained intensive care unit (ICU) settings.

Among the 145 samples evaluated, the majority (90·3%) were classified as pathogenic by the investigator. While this may reflect the predominance of serious infections in ICU patients, our analysis showed that a proportion of organisms (9·7%) did not meet criteria for pathogenicity despite being isolated from clinically ill individuals. The fair agreement (κ = 0·28) between microbiologist and investigator underscores a persistent issue in practice—microbiological positivity does not equate to clinical infection. Data from the VAPrapid2 trial (Hellyer et al., 2015) further support this, highlighting that only 30% of patients were microbiologically confirmed to have VAP despite clinical suspicion, reinforcing the importance of integrating microbiological, clinical, and SOFA score parameters.^10^ Labelle et al. (2010) reported that among patients with healthcare-associated pneumonia (N = 870), approximately 34% had growth of non-pathogenic flora, yet a substantial proportion still received antibiotics, highlighting a mismatch between microbiological findings and clinical decision-making.^5^ Our study provides additional empirical evidence reinforcing the necessity of integrating host and clinical parameters into diagnostic and therapeutic decision-making, not solely relying on microbial isolation.

Our study’s strength lies in correlating organism classification with clinical outcomes. Non-pathogen cases were largely untreated, yet 85·7% recovered uneventfully, and only one unrelated death was observed. This suggests that refraining from antibiotic escalation when non-pathogenic status is appropriately identified is both safe and cost-effective. Conversely, among those classified as pathogens, 68 (51·9%) showed clinical improvement with treatment, while 63 (48·1%) did not respond, of which 48 (76·2%) died. This strong association between investigator classification and treatment response (p < 0·001) supports the algorithm’s validity.

Our findings also reaffirm the clinical value of integrating the SOFA score, symptomatology, and radiologic correlation. Patients with pathogenic organisms were significantly more likely to have features like expectoration, tachypnoea, oxygen requirement, and altered mentation (p < 0 05 for all), in line with classical LRTI presentations. Additionally, a rise in SOFA ≥ 2 within 48 hours was found in 82·4% of pathogen cases vs. 21·4% in non-pathogens (p < 0 001), highlighting its utility as a discriminator. These findings are consistent with the 2016 IDSA/ATS guidelines for Hospital Acquired Pneumonia (HAP)/Ventilator associated Pneumonia (VAP), which emphasize the diagnostic value of clinical features such as new or worsening cough, purulent secretions, leucocytosis, and hypoxia.^3,11,12^

The Multiplex PCR (Biofire® FilmArray)® FilmArray PCR panel, while offering rapid and broad-spectrum pathogen detection—including viruses, atypical, and bacteria—was selectively used in patients with suspected respiratory infections, often in critically ill or diagnostically challenging cases. In our study, Multiplex PCR (Biofire® FilmArray) accounted for 9 7% (n = 14) of the total positive samples and was able to detect a total of 27 organisms, comprising 18 bacteria and 9 viruses, along with clinically relevant antimicrobial resistance genes. Notably, all Multiplex PCR (Biofire® FilmArray)-positive samples were ultimately classified as pathogenic by the stepwise model, likely reflecting the high pre-test probability of infection in these cases. However, Multiplex PCR (Biofire® FilmArray)’s high sensitivity poses a diagnostic dilemma, as it may detect nucleic acids from non-viable organisms or colonizers, particularly in patients with recent or ongoing antibiotic therapy.^7,13^

Nunez et al. (2021) found that each 1-point increase in SOFA score was associated with a 30% increased hazard of 30-day mortality (HR = 1·30; 95% CI: 1·12–1·52; p < 0 001), Raveendra et al. (2020) reported significantly higher SOFA scores in non-survivors at VAP diagnosis (p = 0 005), and Kumar et al. (2010) highlighted the survival benefit of early combination antibiotic therapy in septic shock, reinforcing the role of early clinical-based decisions.^14–16^

The most frequently isolated organisms were *Acinetobacter baumannii* (46·9%), *Klebsiella pneumoniae* (42·1%), and *Pseudomonas aeruginosa* (22·1%), mirroring patterns reported in national ICU surveillance data (ICMR-AMRSN 2020). Their predominance among pathogen-classified cases highlights the model’s strength in identifying clinically relevant, multidrug-resistant organisms.^17^

Colonization factors such as invasive mechanical ventilation and tracheostomy, while more prevalent in pathogen cases, did not reach statistical significance (p = 0·116 and 0·465, respectively). Established literature has linked several host-related and environmental factors to increased risk of airway colonization, including recent hospitalization or antibiotic use, poor oral hygiene due to tobacco use or chronic bedridden status, COPD, prior lower respiratory tract infections, bronchiectasis, and neuromuscular disorders.^18–23^ Intubation with mechanical ventilation increases the risk of bacterial pneumonia by 6–20-fold due to disruption of natural host defences, biofilm formation in endotracheal tubes, and promotion of airway colonization. Opportunistic organisms such as *Acinetobacter baumannii, Pseudomonas aeruginosa*, and *Staphylococcus aureus* can initially colonize the endotracheal tube and subsequently progress to ventilator-associated pneumonia^2,24,25.^ Candida colonization of the respiratory tract is a well-recognized phenomenon in ICU patients. In a multicenter study, Timsit et al. (2019) reported that in patients undergoing mechanical ventilation for more than 4 days, bronchial colonization with *Candida* species was not independently associated with the development of VAP (adjusted cause-specific hazard ratio = 0·98; 95% CI: 0·59–1 65; p = 0·95). However, Azoulay et al. (2006) found that *Candida* colonization of the respiratory tract did not correlate with increased mortality; however, it increased the risk for VAP with Pseudomonas.^26^

Such objective evidence supports the rationale for employing integrative clinico-microbiological approaches like our model to guide targeted antimicrobial use. Preventive strategies—including chlorhexidine oral care, semi-recumbent positioning, and subglottic suctioning—have also been shown to reduce VAP risk by limiting colonization and aspiration of oropharyngeal secretions.^27,28^

Such objective evidence supports the rationale for employing integrative clinico-microbiological approaches like our model to guide targeted antimicrobial use. The study’s structured framework demonstrated alignment with clinical response and mortality patterns, reinforcing its utility in antimicrobial stewardship. Importantly, the model enabled withholding antibiotics in non-pathogen cases without adverse effects—a key stewardship goal. Given the global crisis of antimicrobial resistance, such structured, evidence-based approaches are increasingly essential.

Our study has several limitations. First, it was conducted at a single center over a limited 3-month period, which may limit the generalizability of the findings. Second, although clinical features and SOFA scores were systematically assessed using a structured checklist, there remains the potential for interobserver variability in classification. Third, we did not perform economic or cost analyses, and laboratory inflammatory markers such as C-reactive protein (CRP) and procalcitonin (PCT) were not analysed, which could have further strengthened pathogen differentiation and prognostic assessment; therefore, conclusions regarding resource utilization must be interpreted cautiously. Fourth, the follow-up period was limited to 28 days, and long-term outcomes such as readmissions or recurrent infections were not assessed.

## Conclusion

This prospective study evaluated 145 lower respiratory tract isolates using a structured six-step clinico-microbiological algorithm, classifying 90·3% as pathogens and 9·7% as non-pathogens. The model showed a strong association between classification and clinical outcomes: 85·7% of non-pathogen cases were safely managed without antibiotics, while among pathogen-classified cases, 51·9% improved with treatment and 36·6% succumbed despite therapy. Key predictors of pathogenicity included localized respiratory symptoms, SOFA ≥ 2, and radiologic features. Cohen’s Kappa between the investigator and microbiologist classification was 0·28, indicating fair agreement. This model effectively differentiated infection from colonization and allowed rational antibiotic use without compromising patient safety, reinforcing its potential utility in guiding antimicrobial stewardship efforts in high-burden clinical settings.

## Data Availability

All data produced in the present study are available upon reasonable request to the authors

## Institutional review board statement

The study was reviewed and approved by the Institutional Ethics Committee of All India Institute of Medical Sciences, Rishikesh, India.

## Informed consent statement

All study participants provided informed written consent before enrolling in the study.

## Authors’ contributions

All authors made substantial contributions to conception and design, acquisition of data, or analysis and interpretation of data; took part in drafting the article or revising it critically for important intellectual content; agreed to submit to the current journal; gave final approval of the version to be published; and agree to be accountable for all aspects of the work.

## Acknowledgment

None

## References

1. Averin A, Sato R, Begier E, Yacisin K, Houde L, Lonshteyn A, et al. Short-term and Long-term Mortality Following Hospitalized and Ambulatory Lower Respiratory Tract Illnesses Among US Adults. Open Forum Infect Dis. 2025 Mar 27;12(4).

2. Motallebirad T, Mohammadi MR, Jadidi A, Safarabadi M, Kerami A, Azadi D, et al. Tracheal tube infections in critical care: A narrative review of influencing factors, microbial agents, and mitigation strategies in intensive care unit settings. SAGE Open Med. 2024 Jan 16;12.

3. Kalil AC, Metersky ML, Klompas M, Muscedere J, Sweeney DA, Palmer LB, et al. Management of Adults With Hospital-acquired and Ventilator-associated Pneumonia: 2016 Clinical Practice Guidelines by the Infectious Diseases Society of America and the American Thoracic Society. Clinical Infectious Diseases. 2016 Sep 1;63(5):e61–111.

4. Fabre V, Davis A, Diekema DJ, Granwehr B, Hayden MK, Lowe CF, et al. Principles of diagnostic stewardship: A practical guide from the Society for Healthcare Epidemiology of America Diagnostic Stewardship Task Force. Infect Control Hosp Epidemiol. 2023 Feb 14;44(2):178–85.

5. Labelle AJ, Arnold H, Reichley RM, Micek ST, Kollef MH. A Comparison of Culture-Positive and Culture-Negative Health-Care-Associated Pneumonia. Chest. 2010 May;137(5):1130–7.

6. Dung TTN, Phat VV, Vinh C, Lan NPH, Phuong NLN, Ngan LTQ, et al. Development and validation of multiplex real-time PCR for simultaneous detection of six bacterial pathogens causing lower respiratory tract infections and antimicrobial resistance genes. BMC Infect Dis. 2024 Feb 7;24(1):164.

7. Calderaro A, Buttrini M, Farina B, Montecchini S, De Conto F, Chezzi C. Respiratory Tract Infections and Laboratory Diagnostic Methods: A Review with A Focus on Syndromic Panel-Based Assays. Vol. 10, Microorganisms. MDPI; 2022.

8. A Randomized Trial of Diagnostic Techniques for Ventilator-Associated Pneumonia. New England Journal of Medicine. 2006 Dec 21;355(25):2619–30.

9. Solé Violán J, Fernández JA, Benítez AB, Cendrero JAC, de Castro FR. Impact of quantitative invasive diagnostic techniques in the management and outcome of mechanically ventilated patients with suspected pneumonia. Crit Care Med. 2000 Aug;28(8):2737–41.

10. Hellyer TP, McAuley DF, Walsh TS, Anderson N, Conway Morris A, Singh S, et al. Biomarker-guided antibiotic stewardship in suspected ventilator-associated pneumonia (VAPrapid2): a randomised controlled trial and process evaluation. Lancet Respir Med. 2020 Feb;8(2):182–91.

11. Mandell Lionel A., Niederman Michael S. Harrison’s principles of internal medicine. 21st ed. Loscalzo Joseph, editor. New Delhi: Mc Graw Hill; 2022. 1009–1020 p.

12. Vincent JL, Moreno R, Takala J, Willatts S, De Mendonça A, Bruining H, et al. The SOFA (Sepsis-related Organ Failure Assessment) score to describe organ dysfunction/failure. On behalf of the Working Group on Sepsis-Related Problems of the European Society of Intensive Care Medicine. Intensive Care Med. 1996 Jul;22(7):707–10.

13. Miller JM, Binnicker MJ, Campbell S, Carroll KC, Chapin KC, Gonzalez MD, et al. Guide to Utilization of the Microbiology Laboratory for Diagnosis of Infectious Diseases: 2024 Update by the Infectious Diseases Society of America (IDSA) and the American Society for Microbiology (ASM). Clinical Infectious Diseases. 2024 Mar 5;

14. Ariel Núñez S, Roveda G, Soledad Zárate M, Emmerich M, Teresa Verón M. Ventilator-associated pneumonia in patients on prolonged mechanical ventilation: description, risk factors for mortality, and performance of the SOFA score. Jornal Brasileiro de Pneumologia. 2021 Jun 1;e20200569.

15. R. RK, N. DG, Kodur N, U. CL, K. V. Comparison of clinical parameters with APACHE-II, Sequential Organ Failure Assessment and Clinical Pulmonary Infection Score scores in predicting treatment outcome of patients with ventilator associated pneumonia. International Journal of Advances in Medicine. 2020 Feb 24;7(3):527.

16. Kumar A, Zarychanski R, Light B, Parrillo J, Maki D, Simon D, et al. Early combination antibiotic therapy yields improved survival compared with monotherapy in septic shock: A propensity-matched analysis*. Crit Care Med. 2010 Sep;38(9):1773–85.

17. Annual Report.

18. Lusuardi M, Capelli A, Cerutti CG, Gnemmi I, Zaccaria S, Donner CF. Influence of clinical history on airways bacterial colonization in subjects with chronic tracheostomy. Respir Med. 2000 May;94(5):436–40.

19. Harlid R, Andersson G, Frostell CG, Jörbeck HJ, Ortqvist AB. Respiratory tract colonization and infection in patients with chronic tracheostomy. A one-year study in patients living at home. Am J Respir Crit Care Med. 1996 Jul;154(1):124–9.

20. Cabello H, Torres A, Celis R, El-Ebiary M, Puig de la Bellacasa J, Xaubet A, et al. Bacterial colonization of distal airways in healthy subjects and chronic lung disease: a bronchoscopic study. European Respiratory Journal. 1997 May 1;10(5):1137–44.

21. Lepainteur M, Ogna A, Clair B, Dinh A, Tarragon C, Prigent H, et al. Risk factors for respiratory tract bacterial colonization in adults with neuromuscular or neurological disorders and chronic tracheostomy. Respir Med. 2019 Jun;152:32–6.

22. Scannapieco FA, Papandonatos GD, Dunford RG. Associations Between Oral Conditions and Respiratory Disease in a National Sample Survey Population. Ann Periodontol. 1998 Jul;3(1):251–6.

23. Bonten MJ, Bergmans DC, Ambergen AW, de Leeuw PW, van der Geest S, Stobberingh EE, et al. Risk factors for pneumonia, and colonization of respiratory tract and stomach in mechanically ventilated ICU patients. Am J Respir Crit Care Med. 1996 Nov;154(5):1339–46.

24. Craven DE, Hjalmarson KI. Ventilator-Associated Tracheobronchitis and Pneumonia: Thinking Outside the Box. Clinical Infectious Diseases. 2010 Aug;51(S1):S59–66.

25. Robinson J. Colonization and infection of the respiratory tract: What do we know? Paediatr Child Health. 2004 Jan;9(1):21–4.

26. Azoulay E, Timsit JF, Tafflet M, de Lassence A, Darmon M, Zahar JR, et al. Candida Colonization of the Respiratory Tract and Subsequent Pseudomonas Ventilator-Associated Pneumonia. Chest. 2006 Jan;129(1):110–7.

27. Safdar N, Crnich CJ, Maki DG. The pathogenesis of ventilator-associated pneumonia: its relevance to developing effective strategies for prevention. Respir Care. 2005 Jun;50(6):725–39; discussion 739-41.28. Klompas M, Branson R, Eichenwald EC, Greene LR, Howell MD, Lee G, et al. Strategies to Prevent Ventilator-Associated Pneumonia in Acute Care Hospitals: 2014 Update. Infect Control Hosp Epidemiol. 2014 Aug 10;35(8):915–36.

